# The Influence of Polypharmacy on Type 2 Diabetes Adverse Cardiovascular Outcomes in a Rural Cohort

**DOI:** 10.64898/2026.04.02.26350053

**Authors:** Jennifer W. Li, Lilia A. Crew, Trisha M. Cox, Brenda F. Canine

## Abstract

**Objective:** In this study, we utilized a large-scale clinical database to evaluate the relationship between polypharmacy and adverse outcomes among type 2 diabetes patients in rural Montana to inform strategies that improve adherence, reduce preventable complications, and promote equitable diabetes care in underserved regions.

**Research Design and Methods:** 591 patients from the Big Sky Care Connect Database (BSCC) with type 2 diabetes and medication history were stratified into 3 cohorts based on prescribed number of medications: (1-4 medications, non-polypharmic), (5-9 medications, polypharmic), and (≥10 medications, hyperpolypharmic). Each cohort was examined for Major Adverse Cardiovascular Events (MACE) and Diabetes Complication Severity Index (DCSI). Descriptive statistics, multivariate logistic regressions, linear regression, and Poisson regression analyses were performed.

**Results:** Medication count was associated with male gender (β = -2.1341, p < 0.001). Both medication count (IRR 1.06 per additional medication, p < 0.001) and age (IRR 1.03 per year, p < 0.001) were significant predictors of MACE. Neuropathy and nephropathy prevalence was statistically significant (p < 0.001) across patient cohorts and increased with medication count.

**Conclusions:** A high prevalence of polypharmacy was observed in type 2 diabetic patients in rural Montana. Polypharmacy was found to be a significant predictor of MACE and increased the odds of nephropathy in this study. While disease severity contributes to higher medication counts, these findings highlight the need for medication review and cross-provider management to minimize adverse outcomes.

**Article Highlights:** Why did we undertake this study?

- The study presents the impact of polypharmacy in the management of type 2 diabetes patients in a rural, medically underserved population in Montana

What is the specific question(s) we wanted to answer?

- Does polypharmacy affect patient outcomes in type 2 diabetes?

What did we find?

- Polypharmacy in rural Montana is linked to higher microvascular complication rates.

What are the implications of our findings?

- Medication count is not only a sign of disease burden but is a modifiable risk factor.

**Graphical abstract:** 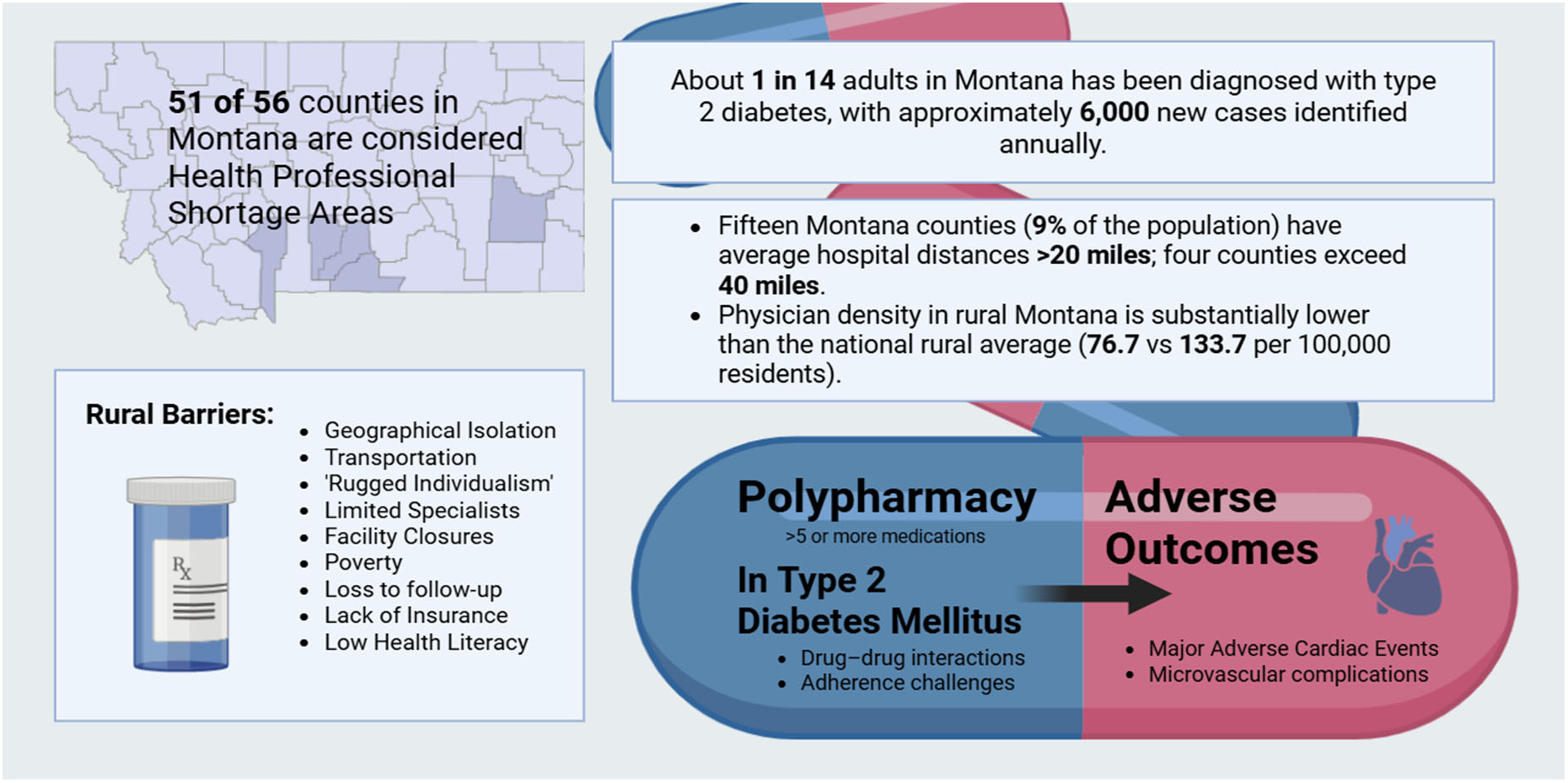

## Introduction

Type 2 Diabetes Mellitus is a chronic, complex disease associated with serious complications affecting numerous body systems, which include heart disease, stroke, amputation, and microvascular disease such as neuropathy, nephropathy, and retinopathy ^1^. Specifically of note, patients with type 2 diabetes are at a higher risk for developing atherosclerotic disease and major adverse cardiac events (MACE) ^2^. Management of type 2 diabetes involves both lifestyle modification and pharmacological treatments aimed at preventing or delaying chronic adverse outcomes and is further complicated by the necessity to treat additional comorbidities.

Polypharmacy, as defined by the National Institute of Health, is the use of five or more medications, and hyperpolypharmacy is the use of ten or more medicationss^3^. Demographic risk factors for polypharmacy include age, chronic conditions, and female sex. Healthcare risk factors include the lack of an established primary care physician, seeing multiple providers, and hospitalizations. The utilization of multiple medications increases the risk of adverse events, as side effect symptoms may be treated with additional medications, resulting in a prescription cascade. With an increasing list of medications, adherence decreases, leading to suboptimal outcomes and heightened risk for complications, mortality, and hospital admissions ^4,5^.

Rural communities face specific systemic and cultural barriers that affect healthcare access and treatment adherence. Despite roughly 20% of the US population living in these rural areas, only about 9% of U.S. physicians work in rural areas, and face a shortage of specialists^6^. Those who seek care may experience difficulties with travel distances and lack of transportation, lack of services, shortages of trained physicians, and internet access ^7,8^. Fragmentations in care, such as patients lost to follow-up, multiple providers, poor record keeping, and automatic refills, also contribute to polypharmacy^9^. Prescribers may be reluctant to interfere with medications prescribed by someone else, and older adults who see the same primary care provider have fewer acute, potentially preventable hospitalizations (PPH)^10^.

In Montana, 51 of the 56 counties are designated as Health Professional Shortage Areas (HPSAs) ^11^. Due to more limited access to resources, individuals in rural areas tend to prioritize self-sufficiency ^12^. This is known as ‘Rugged Individualism’: the belief that one’s identity is rooted in self-sufficiency and self-reliance, and that seeking care may threaten one’s sense of self. This causes medication nonadherence and may cause patients to delay appointments until the disease manifests in a disabling way. This cultural characteristic acts as a key determinant of rural healthcare acceptability^12,13^. Reduced access to healthcare has shown an increased reliance on medication for the management of chronic conditions, and there are limited studies investigating polypharmacy and health outcomes in rural settings (^14^. Approximately 74,400 adults in Montana, or 7.10% of the adult population, have been diagnosed with type 2 diabetes, with an estimated 6,000 additional adults in Montana being diagnosed yearly^15^. Montana is an aging population, with the number of newly diagnosed diabetics expected to increase as the population continues to age ^16^.

Diabetes management in resource-limited settings, such as rural Montana, presents distinct clinical and structural challenges that are often underrepresented in the literature. When access to care is limited, it is also critical to investigate how treatment decisions are made, taking into account cultural practices that influence medication, use, and adherence. Our study of a rural statewide dataset of patients with type 2 diabetes fills the gap to inform practitioners of how resource constraints and treatment complexity intersect to affect patient health.

## Research Design and Methods

This retrospective, observational cohort study was conducted with data from the Big Sky Care Connect Database (BSCC). The BSCC was created in 2018 as a nonprofit organization to provide a statewide coordinated health information exchange (HIE) with the goal of providing data that may improve health outcomes, cost efficiency, and quality of care for all Montanans. The database houses electronic medical record (EMR) data from participants from clinics throughout Montana. The dataset contained complete deidentified patient data from 2020 to 2024.

### Cohort construction

19,809 patients with type 2 diabetes in the BSCC database were identified using ICD diagnosis codes, and cross-validated with at least one HbA1C ≥ 6.5 recorded within the last three years. The index date for the study was defined as the first recorded inpatient or outpatient visit during the observational period from 2020-2024. Patients in the database with any diagnosis of type 1 diabetes mellitus and those who were not prescribed any medication or had a lack of medication history were excluded. A total of 591 patients met the criteria for this study. 4 patients could not be analyzed for microvascular disease due to incomplete data, but were analyzed for MACE incidence. Cohorts were separated by the number of medications to determine the effect of polypharmacy further: Cohort 1 (1-4 medications, non-polypharmic), 2 (5-9 medications, polypharmic), 3 (≥ 10 medications, hyperpolypharmic).

### Definition of Polypharmacy

Polypharmacy was defined as ≥ 5 drugs. Hyperpolypharmacy was defined as taking ≥ 10 drugs. The number of maintenance medications was defined as those prescribed during the observational period from 2020-2024. Prescribed medications for acute conditions were excluded, including. antibiotics, antivirals, and strong pain medications (narcotics, non-OTC pain medications), vitamins and supplements, and other OTC topical use creams were also excluded from medication counts to minimize inconsistency for patients who do not report OTC use.

### Adverse Event Identification

Two categories of adverse events were examined: 1) Major Adverse Cardiovascular Events (MACE) and 2) Diabetes Complication Severity Index (DCSI). MACE is a primary composite endpoint used in many trials, including the evaluation of antidiabetic therapies ^17^. In our investigation, we define MACE to include cardiac arrest, pulmonary embolism, myocardial infarction, and stroke: slightly adapted from the well-known 3-point composite cardiac events, which include cardiovascular death, nonfatal myocardial infarction, or nonfatal stroke ^18^. MACE diagnostic codes utilized are detailed in Supplementary Table S1^19^.

DCSI was utilized to stratify the relative mortality risk of participants, based on their diabetic complications and relevant comorbidities. Classification of diabetes-related complications according to the DCSI scoring system, which assigns severity scores (0–2) across seven domains: retinopathy, neuropathy, cerebrovascular, cardiovascular, peripheral vascular, incidence of severe metabolic abnormalities, and nephropathy. A score of 1 indicates moderate disease manifestations, while a score of 2 reflects advanced or severe pathology. Specific diagnostic entities are listed under each score level, providing a structured framework for quantifying complication burden in clinical and research settings. DCSI is a significant predictor of all-cause mortality in diabetic patients (13). Scores and criteria are detailed in Supplementary Table S2.

### Data Cleaning

From the 591 identified patients, all topical ointments (excluding topical hormones), shampoos, mouthwashes, allergy nasal spray, non-albuterol nasal sprays, stool softeners, lozenges, and ophthalmic medications were removed from the pharmacological prescription count. High-potency analgesics and antibiotics, commonly used during surgical interventions or hospitalizations, were also removed due to insufficient information on the indication for treatment, as it was not possible to distinguish between short-term and long-term use reliably.

### Statistical Analysis

Comorbidities of existing coronary heart disease or heart failure, as well as indicators of more severe diabetic complications, were assessed based on DCSI criteria. We checked the prevalence of microvascular diseases, neuropathy, nephropathy, and retinopathy. Subgroups were combined and analyzed separately as Non-polypharmacy (1-4), Polypharmacy (5-9), and Hyperpolypharmacy (≥ 10) to check if there were additional effects of taking an increased number of medications together. Analyses were conducted using Google Colab (Python), including descriptive statistics, multivariate logistic regressions, linear regression, chi-square, and Poisson regression.

## Research Ethics Statement

Data obtained by investigators was from Big Sky Care Connect. Big Sky Care Connect (BSCC) is a nonprofit formed in 2018 to respond to the need for a statewide health information exchange (HIE) to enhance clinical care in communities across Montana. Data was deidentified of personally identifiable information (PII) in accordance with HIPAA guidelines prior to receipt and analysis and the research team had no access to direct or indirect identifiers, nor any means to re-identify individuals.

## Data and Resource Availability

The data analyzed in the current study are available from the corresponding author upon reasonable request.

## Results

In this analysis of patients with type 2 diabetes from a rural state, we compared clinical outcomes across three medication-based cohorts to assess how varying degrees of polypharmacy relate to adverse events. The following results summarize differences in MACE incidence and DCSI scores across cohorts, highlighting key patterns associated with increasing medication burden in this rural Montana population.

EHR data from Montana residents was obtained through a data request from the Big Sky Care Connect database. Patients with a type 2 diabetes diagnosis from any participating clinic in the state for the last 5 years (2019-2023) were included. From that initial criterion, type 2 diabetes diagnosis was verified with an A1C value of 6.5 or greater during the data period. Further exclusionary criteria were applied, including lack of medication history or no medication, resulting in a pool of 591 patients. Patients were categorized into one of 3 cohorts based on the number of medications prescribed: non-polypharmic (1-4), polypharmic (5-9), and hyperpolypharmic (≥10). Female patients made up a majority of the hyperpolypharmic category (≥10 medications), while male patients held a majority in the lower medication categories (Fig. 1, Supplementary Table S3). Hypertension (HTN) was the most prevalent comorbid condition across all medication burden categories (Supplementary Fig. S1).

**Figure 1.**
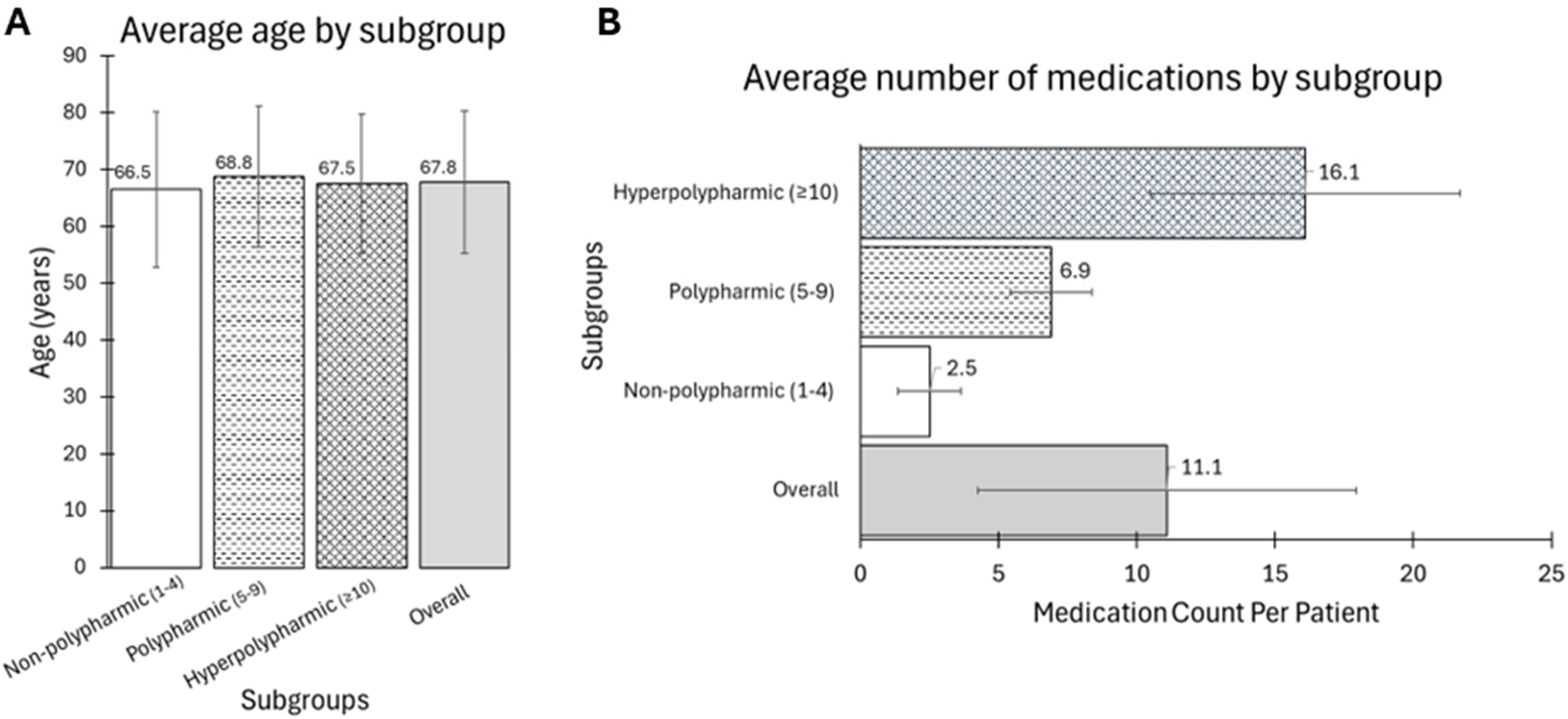
Average Age and Number of Medications by Cohort Medication Subgroup. **(A)** Average participant age of each medication cohort subgroup. **(B)** Average number of medications taken by participants across and within each subgroup.

Diabetes regimen treatments were stratified based on Metformin, insulin, GLP-1 agonists, sulfonylureas, and combinations of other medications (Fig. 2). Metformin-based regimens were shown to be the most common diabetes treatment approach, used by almost 50% of patients, with 19.4% using Metformin monotherapy.

**Figure 2.**
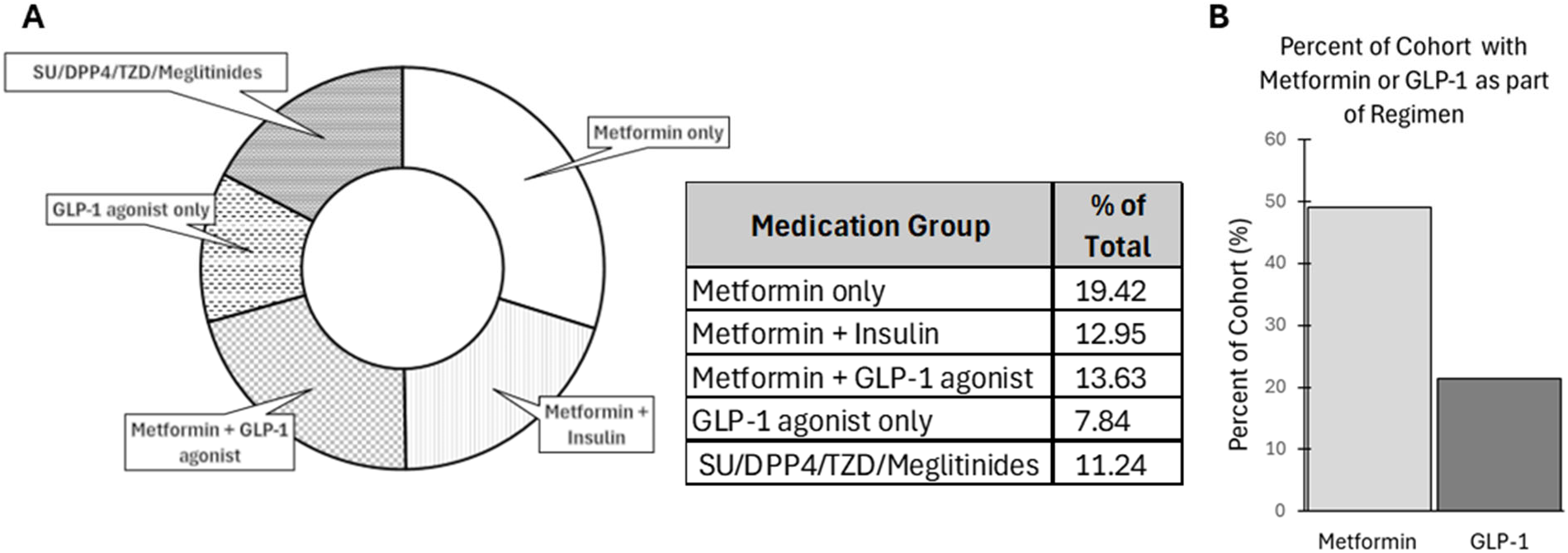
Diabetes Medication Regimens. **(A)** Percentage of diabetes medication regimens across total patients represented as a pie chart and table. **(B)** Percent of the total patient cohort with Metformin and GLP-1 medication either as stand-alone or components of the patients medication regimen.

Patients were assigned a DCSI score based on criteria described in Supplementary Table S2. A multivariable Poisson regression analysis adjusted for DCSI was conducted (Supplementary Fig. S2, Table S4). The model demonstrates a positive association between DCSI and medication count (β = 0.7081, *p* < 0.001), while male gender is inversely associated (β = –2.1341, *p* < 0.001). Age shows a marginal inverse trend (β = –0.0411, *p* = 0.061). Clustering around the identity line suggests reasonable predictive performance. Though age showed a marginally inverse association that did not meet statistical significance criteria (p = 0.061).

A multivariable Poisson regression analysis estimating incidence rate ratios (IRRs) for major adverse cardiovascular events (MACE), controlling for Diabetes Complications Severity Index (DCSI) was examined (Fig. 3, Supplementary Table S5). Independent predictors include age, gender, and medication count. An increasing medication count was independently associated with a higher incidence of MACE (IRR 1.06 per additional medication, 95% CI 1.03-1.08, p < 0.001). Alongside medication count, age was also shown to be a significant predictor of MACE (IRR 1.03 per year, p < 0.001). Male gender was associated with an increased MACE incidence (IRR 1.39, p= 0.0048). DCSI was not a significant predictor in this model.

**Figure 3.**
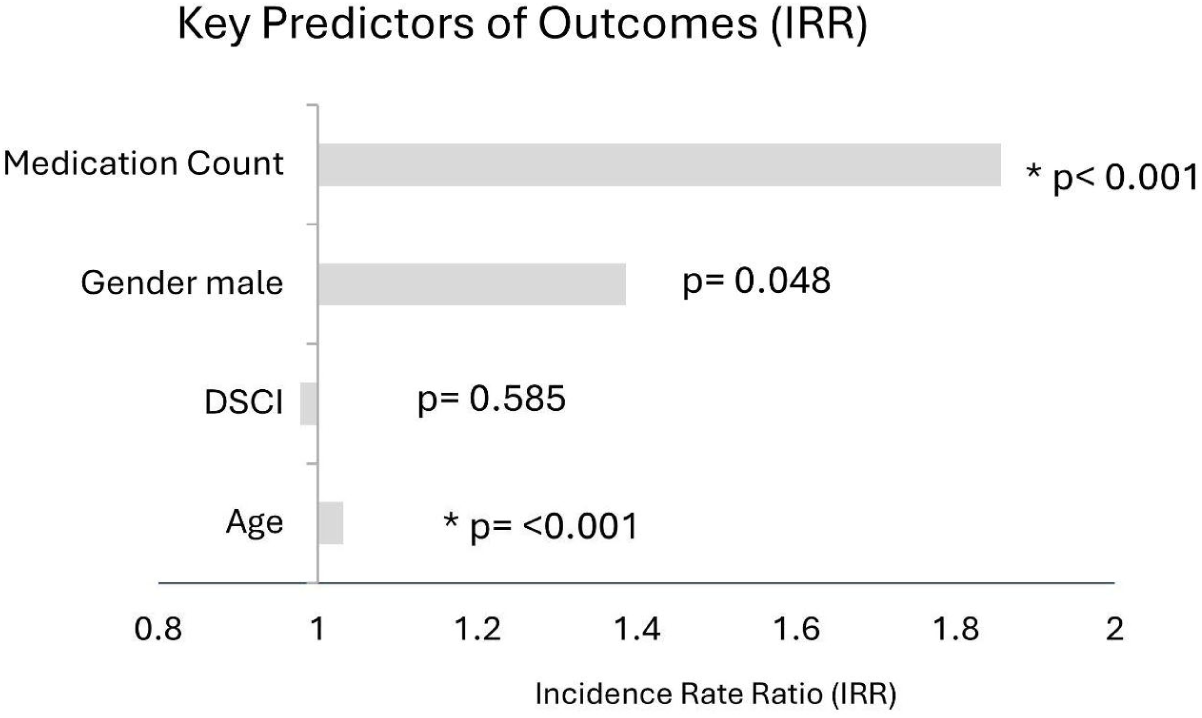
Incidence Rate Ratios of Key Predictors of MACE Outcomes. Multivariate Poisson regression analysis estimating incidence rate ratios (IRRs) for major adverse cardiovascular events (MACE), controlling for Diabetes Complications Severity Index (DCSI). Key predictors tested included medication count, gender (male), DCSI, and age, with reported p-values displayed.

The overall prevalence of neuropathy, nephropathy, and retinopathy was 29.81%, 37.14%, and 8.69%, respectively (Supplementary Table S6). In relation to increased medication burden, neuropathy and nephropathy showed increased incidence (Fig. 4). Chi-square testing revealed statistically significant differences in prevalence for neuropathy and nephropathy across medication categories, while retinopathy did not demonstrate a significant trend (Supplementary Table S6). Neuropathy prevalence rose from 23.81% in patients taking 1-4 medications to 35.8% among those taking ≥ 10 medications (p = 0.0038). As for nephropathy, prevalence rose from 21.43% to 45.6% across the same groups (p < 0.001).

**Figure 4.**
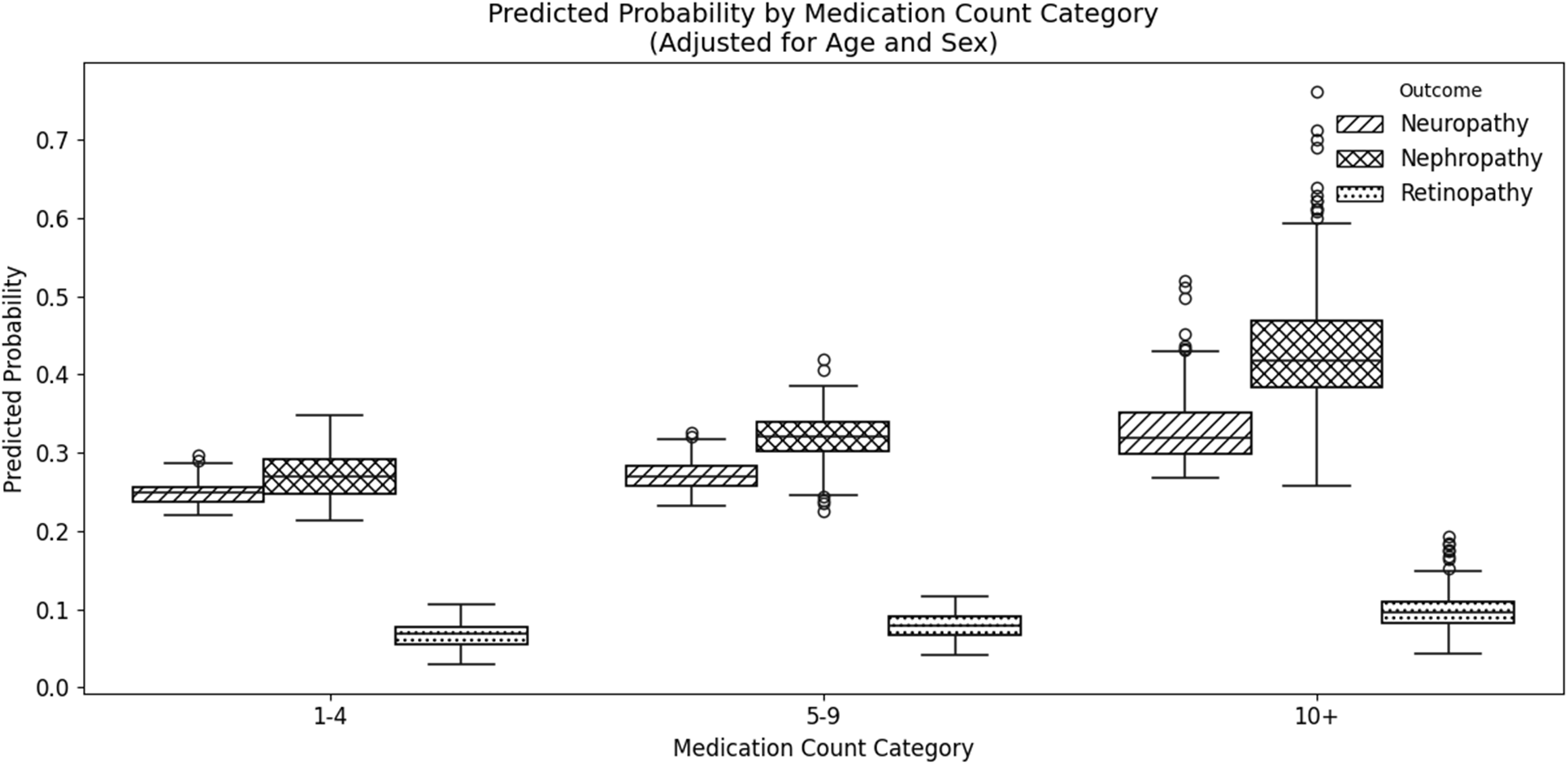
Predicted Probability of Neuropathy, Nephropathy, or Retinopathy Outcomes by Medication Count. Box plots depicting the distribution of predicted probabilities of neuropathy, nephropathy, and retinopathy across increasing medication count categories: 1–4 non-polypharmic, 5–9 polypharmic, and ≥10 hyperpolypharmic.

Multivariate logistic regression models were used to evaluate age, gender, and medication count as predictors of microvascular disease (Supplementary Table S7). Odds ratios and 95% confidence intervals are reported to evaluate effect size and precision. Age and gender were not significant predictors in these models. However, medication count was independently associated with higher odds of neuropathy (OR 1.03 per additional medication, 95% CI 1.00-1.06, p = 0.024) and nephropathy (OR 1.05 per additional medication, 95% CI 1.03-1.08, p < 0.001). No statistically significant association was found between medication count and retinopathy risk (p = 0.132), though the model suggests a potential trend toward increased risk with higher medication burden.

## Conclusion

In this retrospective cohort study of 591 patients with type 2 diabetes from a statewide Montana EMR database, an exceptionally high prevalence of polypharmacy was observed, with patients prescribed an average of 11 medications. 47.38% of the studied population met the criteria for hyperpolypharmacy (≥10 medications) (Supplementary Table S3). As medication count increased, there was a greater percentage of comorbidities, particularly musculoskeletal, respiratory, gastrointestinal, and psychiatric conditions (Supplementary Fig. S1). In contrast, comorbidities such as hypertension, hyperlipidemia, and vascular disease were highly prevalent in all medication categories, though the highest frequencies were observed in the highest medication burden category. As medication count can be used as a proxy for disease, we did not analyze the correlation between these factors.

Males were less likely to be prescribed as many medications in comparison to their female counterparts (Supplementary Table S3). The average age of our cohort was 67.8, with the average age being similar across all subgroups as shown in Fig. 1. In our study, age was only significantly associated with an increased number of MACE events (β=0.0319, p<0.001, Supplementary Table S5), an expected finding. The state of Montana’s aging population (ranked 6th nationally) has many areas designated as Health Professional Shortage Areas, which may also indicate that patients are being seen at a later stage of their diagnosis and have difficulty accessing regular scheduled care. This delay and lack of follow-up may lead to a dependence on prescriptions amongst providers in hopes of reaching patients where they are, as lifestyle interventions are less feasible. Unfortunately, this can lead to over-prescription and medication accumulation with little oversight.

The most common diabetes treatment regimen was Metformin-based (48.73%), with 19.29% of patients receiving Metformin monotherapy (Fig. 2). Men were less likely to be on more prescription medications (β = -2.13, p < 0.001, Supplementary Table S4). As reported, women may have higher medication counts, are likely to be more open to seeking healthcare, and less likely to refuse medication; however adherence may be decreased and warrants further study^20,21^.

Patients with higher DCSI scores were more likely to be on more medications (β = 0.708, p < 0.001) (Fig. 3, Supplementary Table S4). Each additional medication increased the odds of neuropathy (OR = 1.029, CI: 1.0039 - 1.0567) by 2.9% per each additional medication (Fig. 4, Supplementary Table S6). Moreover, each additional medication increased the odds of nephropathy (OR = 1.0527, CI: 1.026- 1.0797) by 5.2% (Supplementary Table S6). There was a non-significant trend of increased retinopathy based on increasing medication number. Age and gender, while expected to be risk factors for microvascular disease, were not significant; only medication count was a statistically significant risk factor for nephropathy and neuropathy (Supplementary Table S7).

Poor glycemic control in diabetic patients has long been implicated in damage to the retina, leading to vision loss, glomeruli leading to kidney dysfunction, and nerve damage leading to neuropathy. In this unique population, polypharmacy was the strongest predictor of diabetic complications (DCSI, MACE) and microvascular disease (nephropathy, neuropathy). In this study, a higher DCSI score was significantly correlated with increased prescriptions, showing a higher disease burden. DCSI was mainly determined from the observable complications of diabetes, and only factors in comorbidities from closely related systems, such as cardiac and renal disease, meaning other factors may be at play, but were not captured in this analysis. patients. Other factors could include other comorbidities such as hypertension, hyperlipidemia, and obesity. It is well established that type 2 diabetes also increases the risk of cardiac disease through inflammation, oxidative stress, and atherosclerosis mechanisms, all of which may be managed through medication, potentially creating prescription cascades and physiological feedback loops, causing even greater medication numbers after the diagnosis of microvascular disease.

Patients who are reluctant to seek medical care and those who are living in an area where screening may not be readily accessible are more likely to forgo diabetic eye screening altogether. In Montana, 15 counties, representing about 9% of the population, have average hospital distances exceeding 20 miles, and in four of those counties, the average distance is more than 40 miles. Rural counties in Montana have 76.7 physicians per 100,000 residents, compared with 233.3 in the state’s non-rural counties and 133.7 in rural counties nationwide ^22^. In fact, Montana only has 103 registered providers of ophthalmology for a state that is 147,000 square miles in size.

This study’s strengths include the use of a large, state-wide EMR dataset representative of rural healthcare in Montana in connection with the application of a Diabetes Complications Severity Index to control for baseline disease burden. The limitations of this study include unmeasured comorbidities, lack of knowledge on medication adherence, and unavailable mortality data. Another note is that providers may not have removed the record of medications a patient is not actively taking or discontinued old medications. One patient had 43 medications still active when we obtained the data from the database, which would pose an enormous burden on the patient. Also, variations in screenings may have led to an underestimation of some outcomes. They also show the real-life effects of low healthcare follow-up, as a significant number of participants were removed due to missing HbA1C values within the last 3 years, when standard care recommends repeat HbA1C every 6 months. Despite the noted limitations, the robust and consistent association between medication count and microvascular outcomes is still noteworthy.

The findings provide further support that polypharmacy is a critical risk factor in the management of diabetes, supporting previous research that has shown that concurrent prescriptions increase adverse drug events, hospitalization, and mortality among patients with chronic conditions (13). This project extends these observations and quantifies the relationship between medication count and microvascular complications in a rural setting, finding that each additional prescribed medication increased the odds of neuropathy and nephropathy. While disease severity contributes to compounding medication counts, these findings highlight the need for medication review and cross-provider management to minimize adverse outcomes. Providers should exercise caution before prescribing additional medications, as they may contribute to drug-drug interactions, altered pharmacokinetics, and reduced treatment adherence, all leading to potentially worse patient outcomes. The results demonstrate the challenges of diabetes care in rural, underserved populations where limited access and cultural factors can further expose medication burdens. Addressing the root issues by completing medication list reviews, patient education, and improved follow-up can be a promising step towards less adverse outcomes and improved quality of life for patients. Future studies should explore causality, medication adherence, and intervention strategies to optimize safe prescribing practices and improve patient-centered outcomes.

## Personal Thanks

The authors would like to give personal thanks to the Weissman Hood Institute Staff and Faculty and Touro University College of Osteopathic Medicine, Montana Campus Office of Research, for helpful discussions and advice during the development of this manuscript.

## Funding and Assistance

This study was supported by the National Institutes of Health (P2GM152335) and Weissman Hood Institute Institutional Funds.

## Author Contributions and Guarantor Statement

J.W.L. conceived and designed the study, collected data, analyzed data, drafted, and edited the manuscript. L.A.C. contributed to data collection, analysis, drafted, and edited the manuscript. T.M.C drafted and edited the manuscript. B.F.C. supervised the project, provided conceptual guidance, secured funding, prepared figures, and critically revised the manuscript for important intellectual content. The authors accept full responsibility for the integrity of the work as a whole, including the accuracy and completeness of the data and analyses. All authors approved the final manuscript and agree to be accountable for all aspects of the work

## Prior Presentation

Data from this manuscript was previously presented on March 10 and April 9, 2025, at Touro COM Research Day 2025 and the Confluence Public Health Alliance.

## Supplemental Material

**Table S1.** ICD-10 Definition of Major Adverse Cardiovascular Events (MACE). This table outlines diagnostic codes relevant to MACE, including acute myocardial infarction subtypes, cardiac arrest, pulmonary complications, and cerebral infarction etiologies. These classifications facilitate standardized documentation, retrospective cohort identification, and outcome tracking in cardiovascular and critical care research. Cardiovascular death was not included in the definition of MACE, as data for all-cause mortality were unavailable.

| Category | ICD | Diagnosis Description |
| --- | --- | --- |
| Myocardial Infarction | I21.4 | Non-ST elevation (NSTEMI) myocardial infarction |
|  | I21.11 | ST elevation (STEMI) myocardial infarction involving right coronary artery |
|  | I21.3 | ST elevation (STEMI) myocardial infarction of unspecified site |
|  | I21.A1 | Myocardial infarction type 2 |
|  | I21.19 | ST elevation (STEMI) myocardial infarction involving other coronary arteries of inferior wall |
|  | I21.02 | ST elevation (STEMI) myocardial infarction involving left anterior descending coronary artery |
|  | I21.9 | Acute myocardial infarction, unspecified |
|  | I22.9 | Subsequent ST elevation (STEMI) myocardial infarction of unspecified site |
| Cardiac Arrest | I46.9 | Cardiac arrest, cause unspecified |
| Pulmonary Events | I26.99 | Other pulmonary embolism without acute cor pulmonale |
|  | I26.02 | Saddle embolus of pulmonary artery with acute cor pulmonale |
|  | I27.82 | Chronic pulmonary embolism |
| Cerebral Events | G45.9 | Transient cerebral ischemic attack, unspecified |
|  | I63.9 | Cerebral infarction, unspecified |
|  | I63.10 | Cerebral infarction due to embolism of unspecified precerebral artery |
|  | I63.11 | Cerebral infarction due to embolism of left middle cerebral artery |
|  | I63.11 | Cerebral infarction due to embolism of right middle cerebral artery |
|  | I63.511 | Cerebral infarction due to unspecified occlusion or stenosis of right middle cerebral artery |
|  | I63.512 | Cerebral infarction due to unspecified occlusion or stenosis of left middle cerebral artery |
|  | I63.432 | Cerebral infarction due to embolism of left posterior cerebral artery |
|  | I63.441 | Cerebral infarction due to embolism of right cerebellar artery |

**Table S2.**
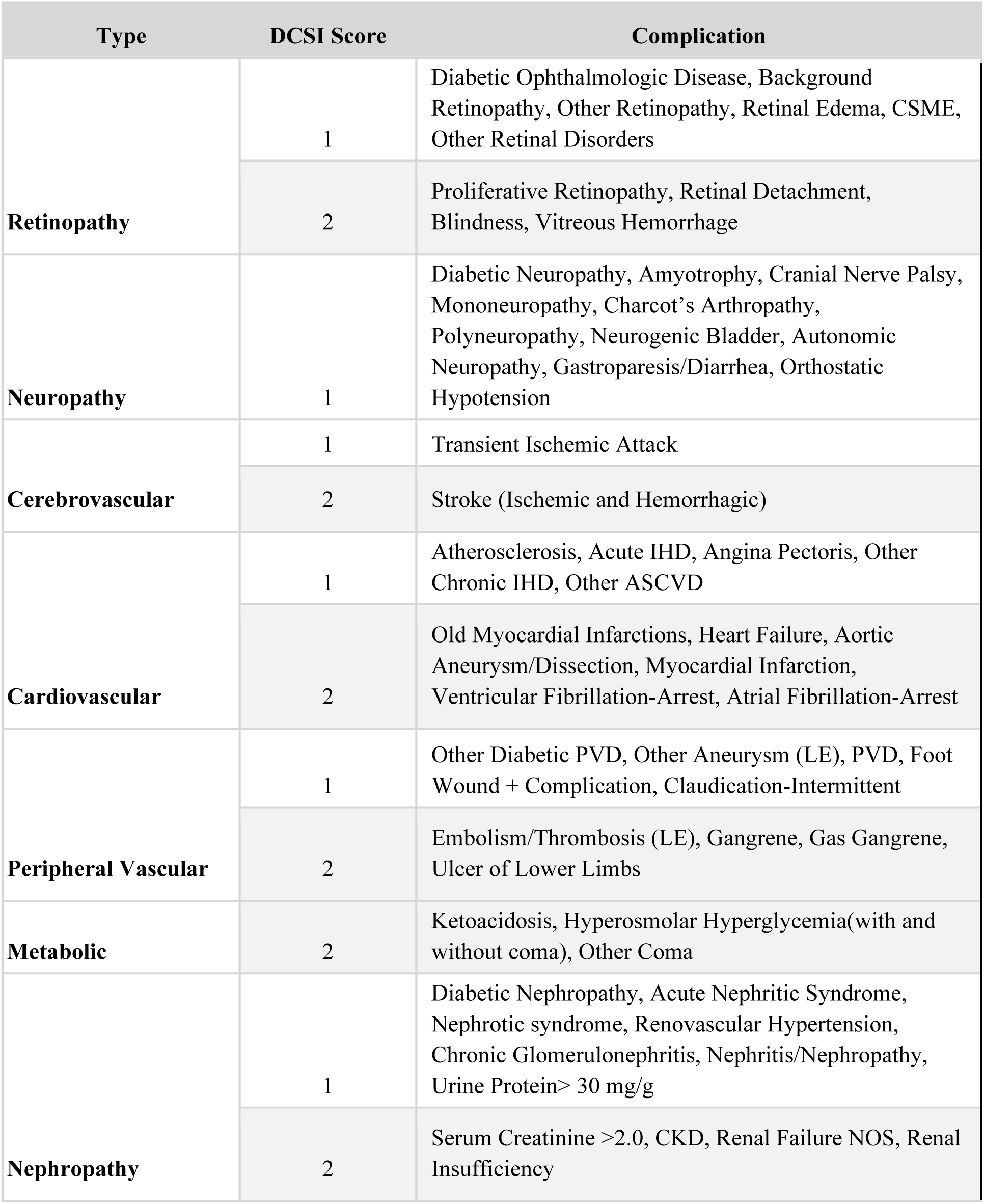
Diabetes Complications Severity Index (DCSI) Scoring by Complication Type. Classification of diabetes-related complications according to the DCSI scoring system, which assigns severity scores (0–2) across seven domains: retinopathy, neuropathy, cerebrovascular, cardiovascular, peripheral vascular, incidence of severe metabolic abnormalities, and nephropathy. A score of 1 indicates moderate disease manifestations, while a score of 2 reflects advanced or severe pathology.

**Table S3.** Participant Counts Across Stratified Age Groups and Gender Categories. Descriptive statistics summarizing the number of medications taken by participants across stratified age groups and gender categories. Columns represent medication burden ranges from 1–4, 5-9 to ≥10 medications, with corresponding participant counts and mean ± SD values.

|  | Overall | 1 to 4<br>Non-polypharmic | 5 to 9<br>Polypharmic | ≥10<br>Hyperpolypharmic |
| --- | --- | --- | --- | --- |
| # of Participants | 591 | 84 | 198 | 309 |
| # of Medications | 11.142 ± 6.845 | 2.583±1.143 | 6.980±1.474 | 16.136±5.660 |
| Age | 67.831±12.522 | 66.547±13.691 | 68.848±12.381 | 67.528±12.271 |
| <30 | 0.338 | 1.190 | 0.000 | 0.324 |
| 30-39 | 2.030 | 3.571 | 2.525 | 1.294 |
| 40-49 | 7.107 | 8.333 | 6.566 | 7.120 |
| 50-59 | 14.552 | 16.667 | 10.101 | 16.828 |
| 60-69 | 23.858 | 20.238 | 26.768 | 22.977 |
| 70-79 | 34.687 | 32.143 | 34.343 | 35.599 |
| >80 | 17.428 | 17.857 | 19.697 | 15.858 |
| Male | 47.885 | 54.762 | 55.051 | 41.424 |
| Female | 51.438 | 45.238 | 43.939 | 57.929 |
| Unknown | 0.677 | 0.000 | 1.010 | 0.647 |

**Table S4.** Multivariate Linear Regression Analysis of Key Predictors of Number of Medications Prescribed. Regression model estimating the number of medications prescribed as a function of adjusted Diabetes Complications Severity Index (DCSI), age, and gender.

| Variable | Regression Coefficient ($\beta$) | Std. Error | t-value | p-value | 95% CI ($\beta$) |
| --- | --- | --- | --- | --- | --- |
| Constant | 13.4221 | 1.536 | 8.738 | <0.001 | [10.405, 16.439] |
| DCSI | 0.7082 | 0.132 | 5.355 | <0.001 | [0.448, 0.968] |
| Age | -0.0411 | 0.022 | -1.874 | 0.061 | [-0.084, 0.002] |
| Gender (male) | -2.1341 | 0.548 | -3.897 | <0.001 | [-3.210, -1.058] |

**Table S5.** Poisson Regression Modeling of MACE Incidence Adjusted for DSCI. Multivariable Poisson regression results estimating incidence rate ratios (IRRs) for major adverse cardiovascular events (MACE), controlling for Diabetes Complications Severity Index (DCSI). Independent predictors include age, gender, and medication count.

| Variable | Coefficient ($\beta$) | Std. Error | z-value | p-value | 95% CI ($\beta$) | IRR | 95% CI (IRR) |
| --- | --- | --- | --- | --- | --- | --- | --- |
| Constant | -4.3789 | 0.576 | -7.601 | <0.001 | [-5.508, -3.250] | 0.0125 | [0.0041, 0.0388] |
| Age | 0.0319 | 0.007 | 4.294 | <0.001 | [0.017, 0.046] | 1.0324 | [1.0175, 1.0476] |
| DCSI | -0.0218 | 0.040 | -0.547 | 0.585 | [-0.100, 0.056] | 0.9784 | [0.9048, 1.0581] |
| Gender (male) | 0.3274 | 0.166 | 1.976 | 0.048 | [0.003, 0.652] | 1.3873 | [1.0026, 1.9200] |
| Medication Count | 0.0550 | 0.011 | 5.125 | <0.001 | [0.034, 0.076] | 1.0565 | [1.0345, 1.0790] |

**Table S6.**
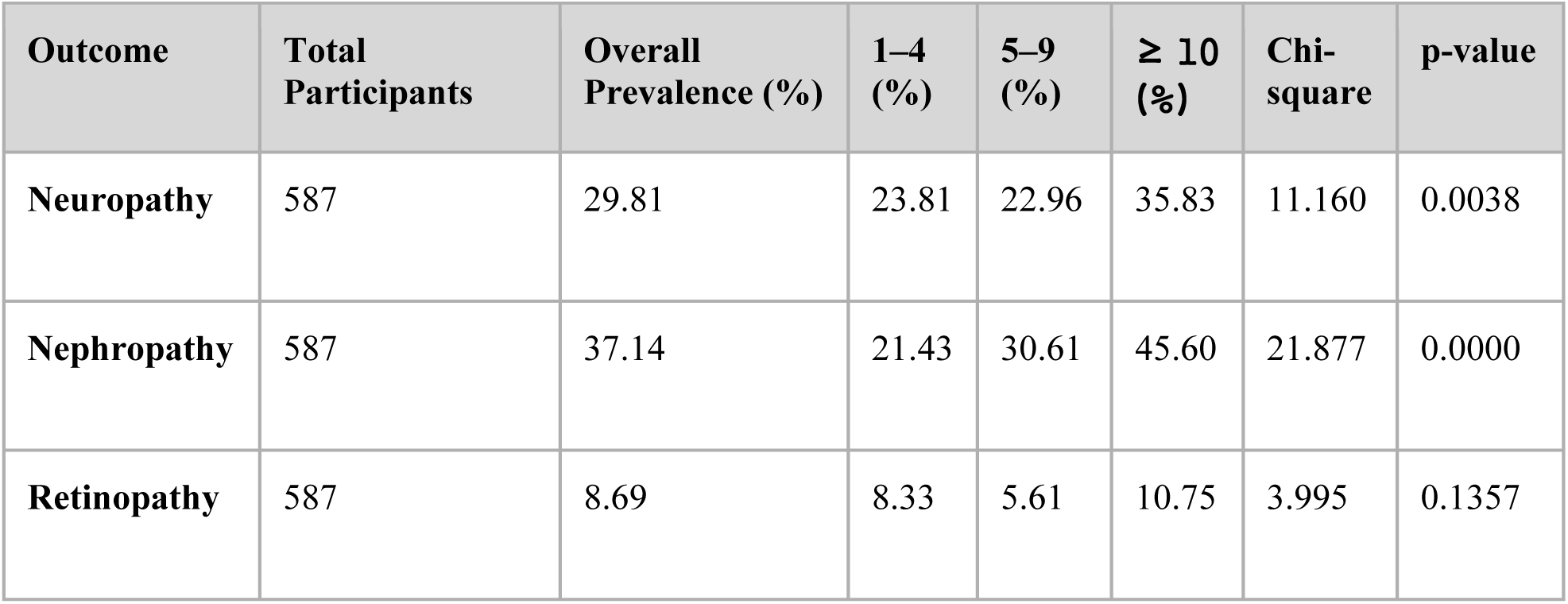
Neuropathy, Nephropathy, and Retinopathy Overall Prevalence. Chi-square significance testing of overall presence of neuropathy, nephropathy, and retinopathy across medication counts.

| Outcome | Total Participants | Overall Prevalence (%) | 1-4 (%) | 5-9 (%) | ≥ 10 (%) | Chi-square | p-value |
| --- | --- | --- | --- | --- | --- | --- | --- |
| Neuropathy | 587 | 29.81 | 23.81 | 22.96 | 35.83 | 11.160 | 0.0038 |
| Nephropathy | 587 | 37.14 | 21.43 | 30.61 | 45.60 | 21.877 | 0.0000 |
| Retinopathy | 587 | 8.69 | 8.33 | 5.61 | 10.75 | 3.995 | 0.1357 |

**Table S7.** Neuropathy, Nephropathy, and Retinopathy Risk Factors. Multivariable logistic regression modeling of neuropathy, nephropathy, and retinopathy predictors: age, medication count, male gender at birth, and constant.

|  | Neuropathy |  | Nephropathy |  | Retinopathy |  |
| --- | --- | --- | --- | --- | --- | --- |
| Variable | OR [95% CI] | p-value | OR [95% CI] | p-value | OR [95% CI] | p-value |
| Age | 0.99 [0.98-1.01] | 0.449 | 1.01 [1.0-1.02] | 0.195 | 1.02 [0.99-1.04] | 0.156 |
| Medication Count | 1.03 [1.0-1.06] | 0.024 | 1.05 [1.03-1.08] | 0.000 | 1.03 [0.99-1.07] | 0.132 |
| Gender (Male) | 1.06 [0.74-1.53] | 0.736 | 0.87 [0.62-1.23] | 0.434 | 1.19 [0.66-2.14] | 0.558 |
| Constant | 0.43 [0.15-1.22] | 0.113 | 0.19 [0.07-0.53] | 0.002 | 0.02 [0.0-0.12] | 0.000 |

**Figure S1.**
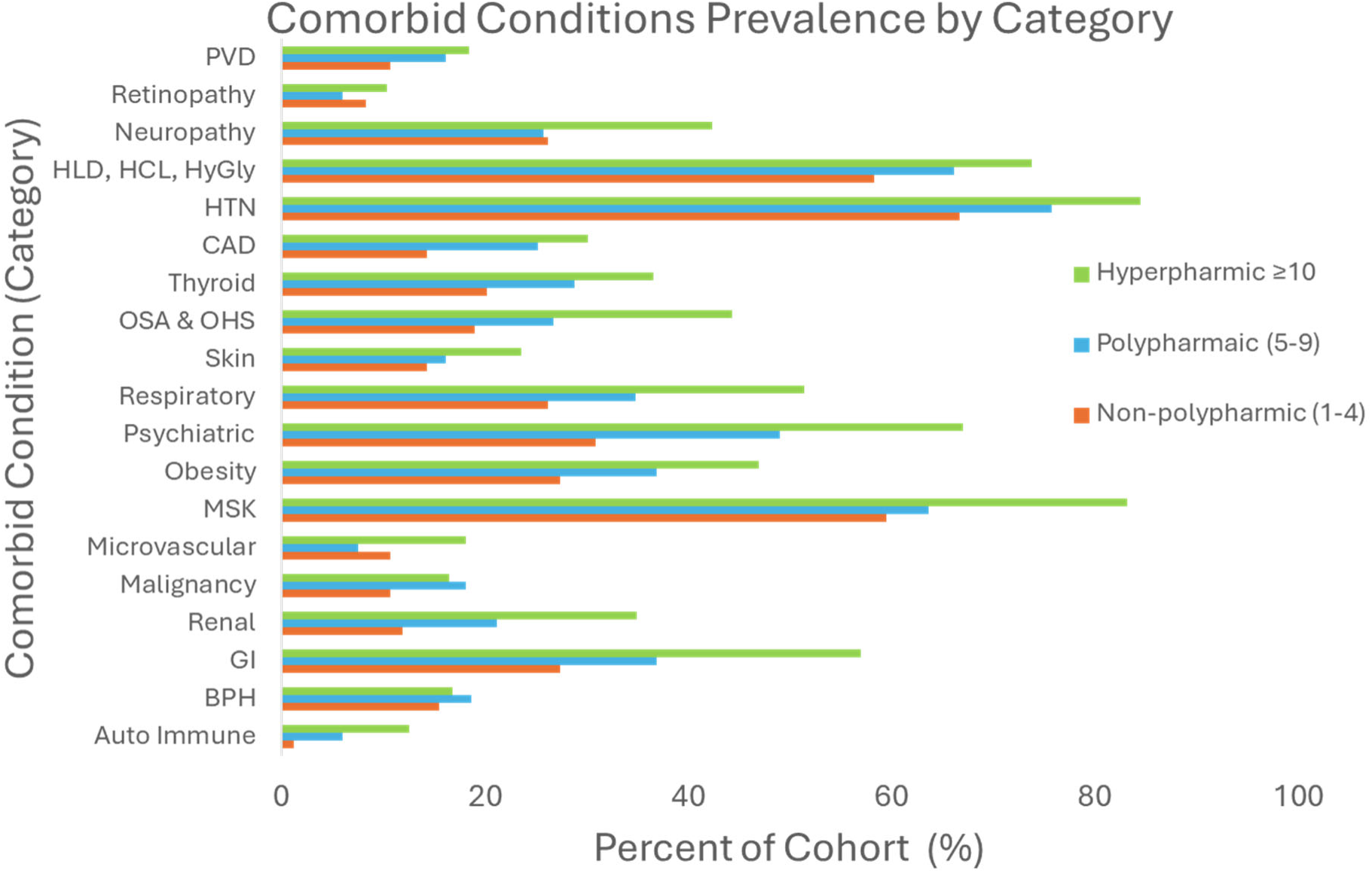
Comorbid Conditions of Participants. Prevalence of comorbid conditions represented as percent of cohort stratified by medication burden ranges: non-polypharmic, polypharmic, and hyperpolypharmic.

**Figure S2.**
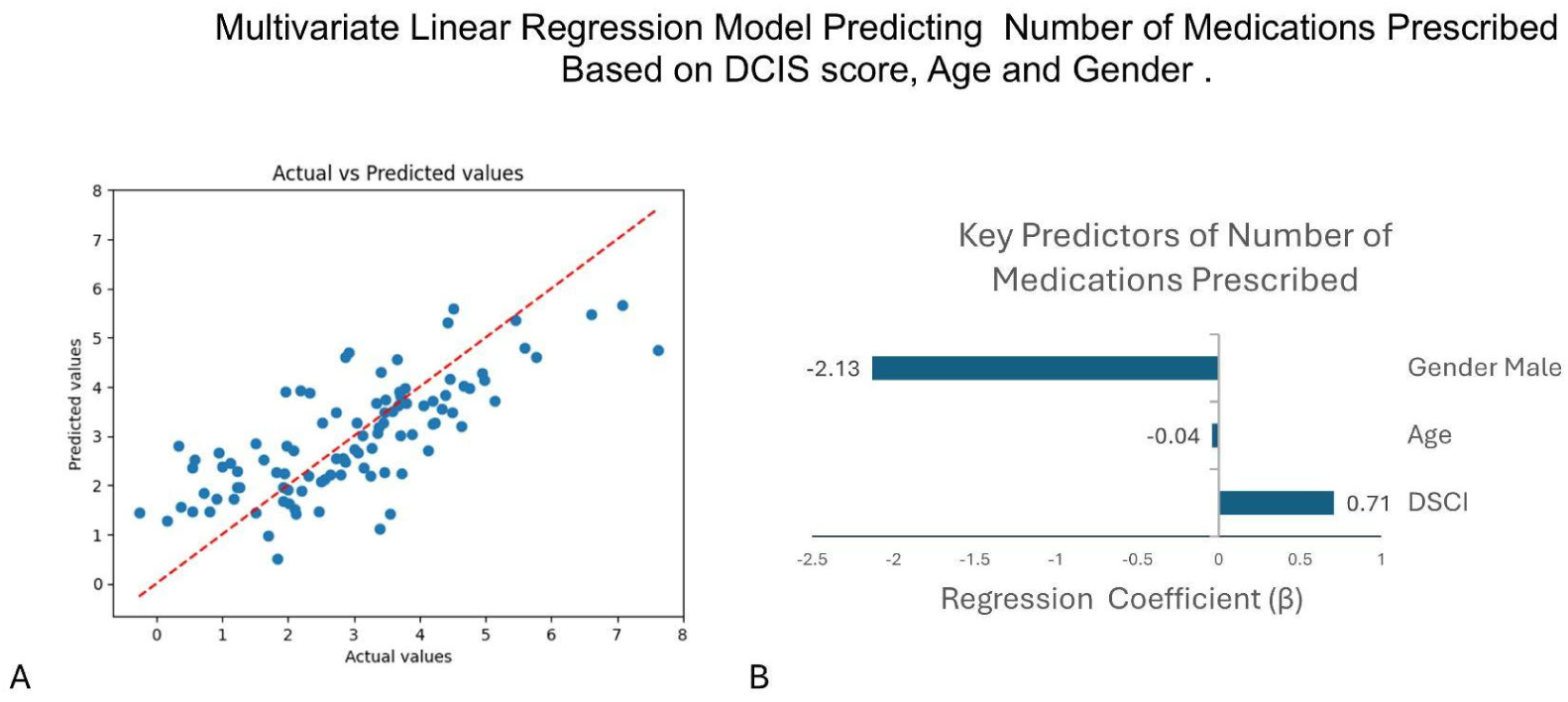
Actual vs. Predicted Number of Medications Prescribed and Key Predictors of Amount of Medications Prescribed Based on Multivariate Linear Regression Analysis. **(A)** Scatter plot comparing observed versus predicted values for the number of medications prescribed, derived from a multivariate linear regression model incorporating DCSI score, age, and gender. **(B)** Regression coefficients (ꞵ) of medication count association with gender (male), age, and DSCI.

